# Predicting clinical outcomes and hospitalization stay of hospitalized COVID-19 patients by using Deep Learning methods

**DOI:** 10.1101/2022.01.28.22270040

**Authors:** Arturas Ziemys

## Abstract

Predicting outcomes and other critical clinical events of hospitalized COVID-19 patients may provide a valuable asset to healthcare and a chance to improve patient outcomes. Here, we have analyzed over 10,000 hospitalized COVID-19 patients in the Houston Methodist Hospital at the Texas Medical Center from the beginning of pandemics till April of 2020. This work extends our previous study analyzing longitudinal symptomatics of the hospitalized patients by seeking to understand how standard patient clinical data, like demographics and comorbidities, together with symptom data from early hospitalization can be used to predict the clinical outcomes and hospitalization stay. Deep Learning (DL) classification and regression methods were applied to quantify patient record importance and to perform predictions. The results suggest that patient outcome can be predicted with up to 75% accuracy. However, the prediction of hospitalization stay was more complex indicating deeper optimization of features.

## Introduction

…

There is large body of studies and knowledge dedicated to COVID 19 infection and patient outcomes. However, the most of them do not embark on the potentially valuable information source of longitudinal symptomatic. Here, we seek to understand the importance of longitudinal symptomatics in the prediction of clinical outcomes and hospitalization stay by using DL methods. This study capitalizes on the findings of our previous study about longitudinal symptomatic [1] and approaches the clinical data set from the large data and AI perspective.

## Material and methods

### Data source

The study protocol was reviewed by the COVID Retrospective Research Task Force and due to the de-identified nature of the data set used, the study was granted a waiver from the IRB. The deidentified data set was acquired from the CURATOR data base in the HMH (PRO00025445). Only adult patients tested positive for COVID-19 were included. The ages of patients older than 90 year were capped at 90 years for deidentification purposes. Time records associated with patient were offset by the admission time.

### Symptom anatomical aggregation

Approximately 40 unique symptoms were extracted from records originating from patient flowsheets. To make the analysis more robust, we have aggregates symptoms based on their anatomical associations. Table 1 presents symptom groups, their abbreviations, and unique symptoms attributed to symptom groups. The symptom grouping is unique so that each individual symptom belongs to one symptom group only. The stiff neck symptom was attributed to the Central Nervous System (CNS) group because other viral brain infections, like viral meningitis [2, 3], possess such a symptom. Eight unique symptom groups were created. Symptom remission was defined as a rate of symptom frequency change over time within a specific group.

### Preprocessing

All patients records missing clinical variables were excluded from analysis. Because the data set contains ∼:10 ratio between deceased and alive patients, the data set was transformed into the 2000 patients data sets having 1:1 ratio between deceased and alive patients. Collinearity analysis in feature selection was used to remove correlated variables (Figure 1). The dominant number of features were poorly correlated justifying their use in the model development.

**Figure 1.**
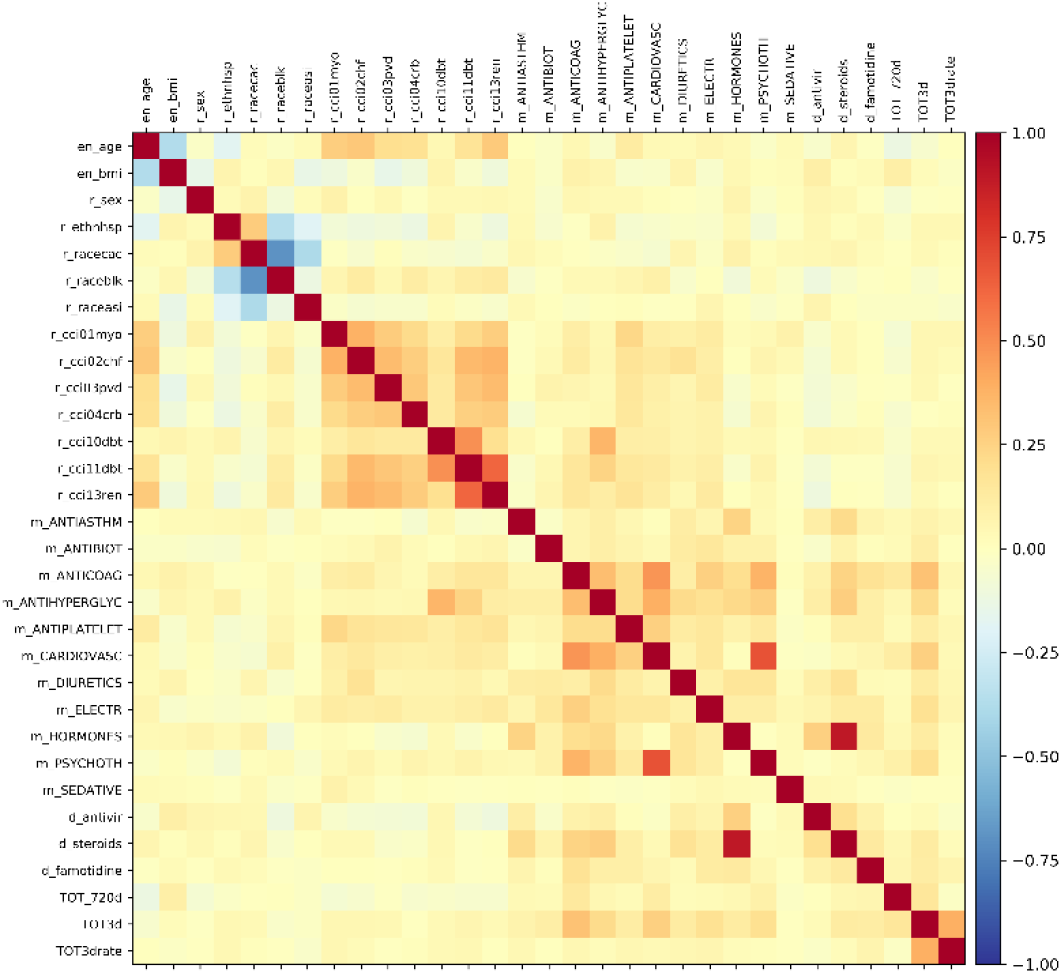
Feature cross-correlation by suing Pearson correlation coefficient R ranging from -1 to 1 (the color bar)

### Deep neural network and feature selection

DL models were implemented by using the Keras [4] and TensorFlow [5] framework executed in Python 3. Rectified linear units were chosen as activation functions. The output layer used a sigmoid function for binary classification. Five-fold repeated random sub-sampling validation for model cross-validations. Optimal model search was conducted by using randomized models within provided ranges: batch size – 4 – 128, number of hidden layers – 1 – 10, number of nodes – 1x – 3x of number of features. Regularization was also randomized with dropout rate decrease from first hidden layer (upper value 0.4) to the last hidden layer (lower value 0.05). Classification models were complied with the binary cross-entropy loss function and the steepest gradient descent optimizer, while regression models were compiled with the mean square error as a loss function and Adam optimizer. The importance of features was scored by using permutations approach and sorter for further analysis. The optimal number of features was derived by performing manual was derived through comparing model accuracy of testing with different numbers of features.

## Results and discussion

### Patient classification based on outcomes

The permutation analysis has revealed that patient age, administrated electrolytes, steroids, and comorbidities like have the most influence to the model (Figure 2). The collinearity of the top 11 features was low, except among few comorbidities (R 007E 0.5). All top features were used for outcome model optimization. We have performed randomized parameter optimization for the classification model by performing 1000 random models with 5-fold repeated random sub-sampling validation for model cross-validations. The average overall model accuracy of the top five models was 0.727±0.001. The prediction scores for alive and deceased patient were the following: precision - 0.768±0.007 and 0.698±0.005, recall – 0.657±0.022 and 0.798±0.019, and F1 – 0.707±0.009 and 0.744±0.007. The performance of the best model is illustrated in the Figure 3 by showing the corresponding ROC curve, history of loss over the curse of epochs, and the confusion matrix. The results indicate significant number of true positive outcomes among patients.

**Figure 2.**
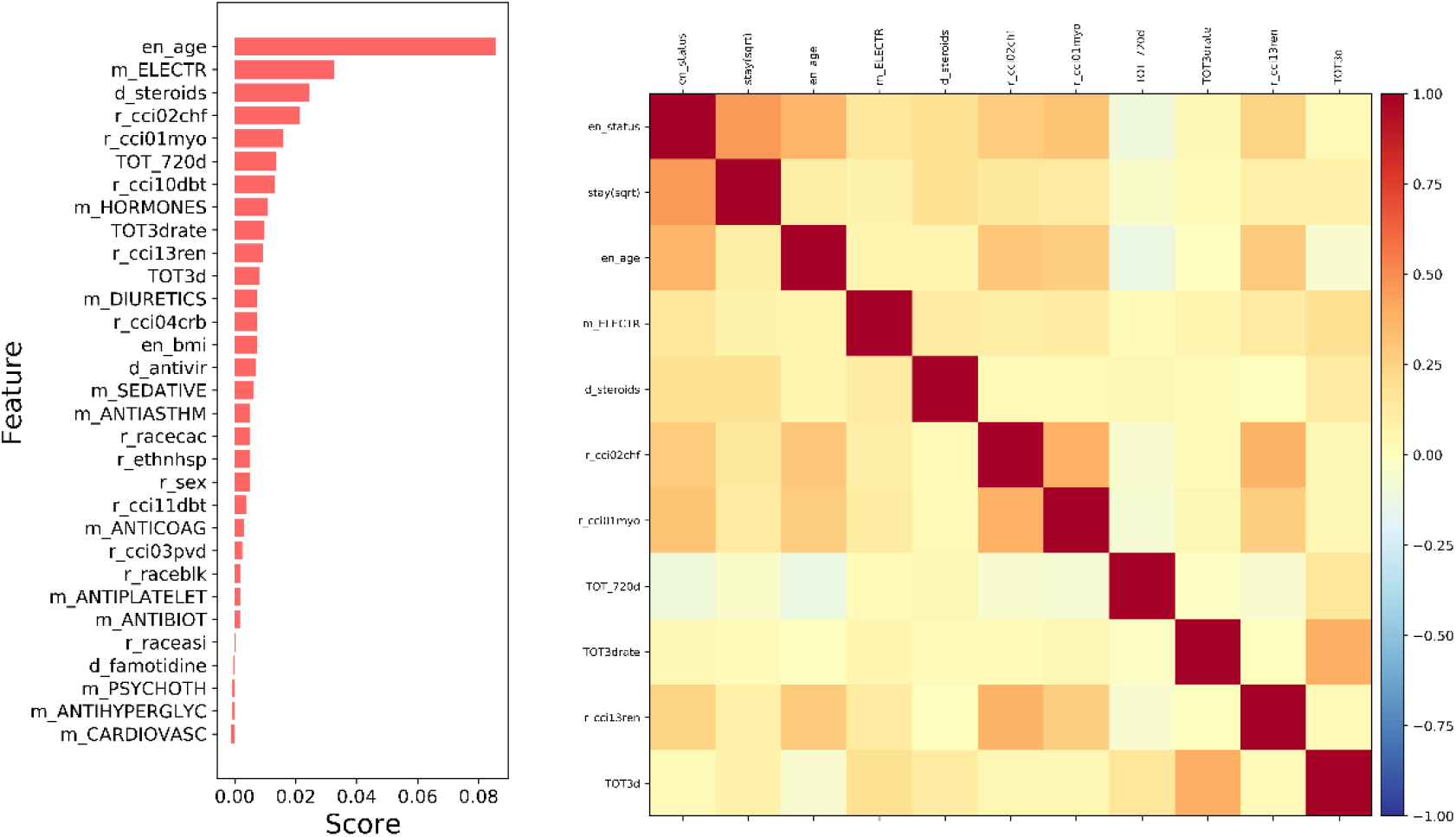
Features scoring for outcomes classification. (*left*) Permutation feature scoring. (*right*) Cross-correlation by suing Pearson correlation coefficient R ranging from -1 to 1 (the color bar).

**Figure 3.**
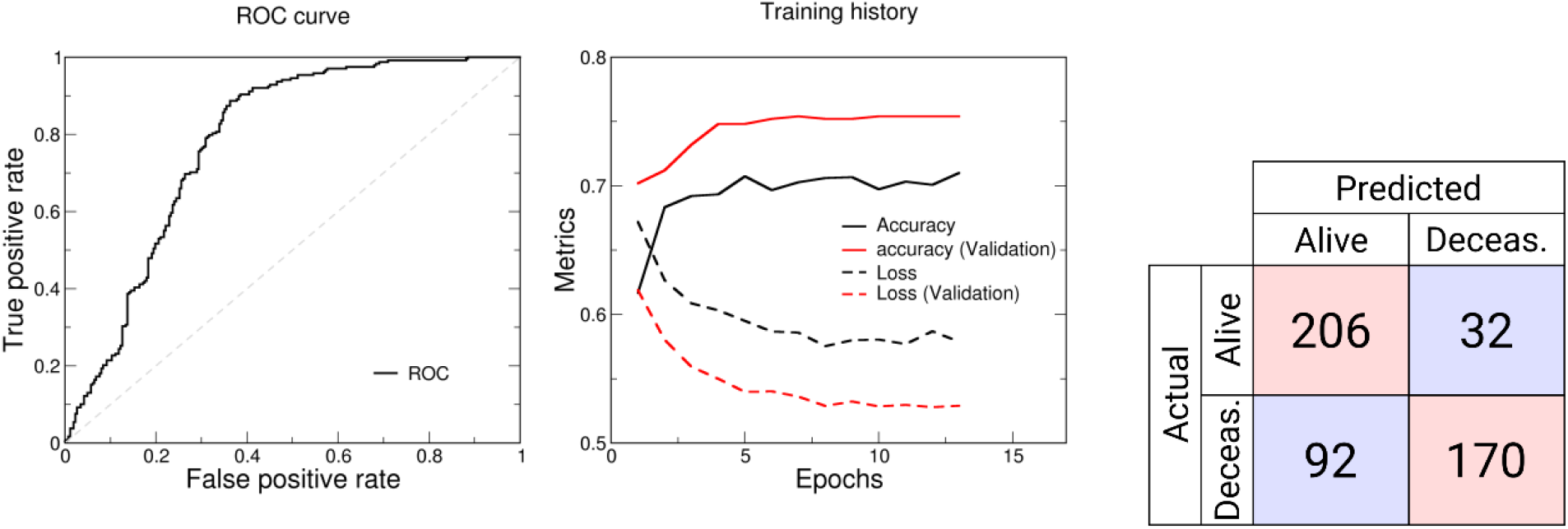
Patient outcome prediction by DL classification model. (*left*) ROC curve and training history of the best classification model. (*right*) The confusion matrix.

### Hospitalization stay prediction by DL regression model

The feature scoring results were slightly different compared to features scored by the outcomes model (Figure 4). Medication and BMI was found more important. The symptom remission rate at the first hospitalization days was also found important along with cardiovascular and renal comorbidities. The model optimization resulted in overfitting and poor prediction, which indicated deeper analysis and optimization is needed to optimize the set of feature, the size of the set, and other DL model parameters.

**Figure 4.**
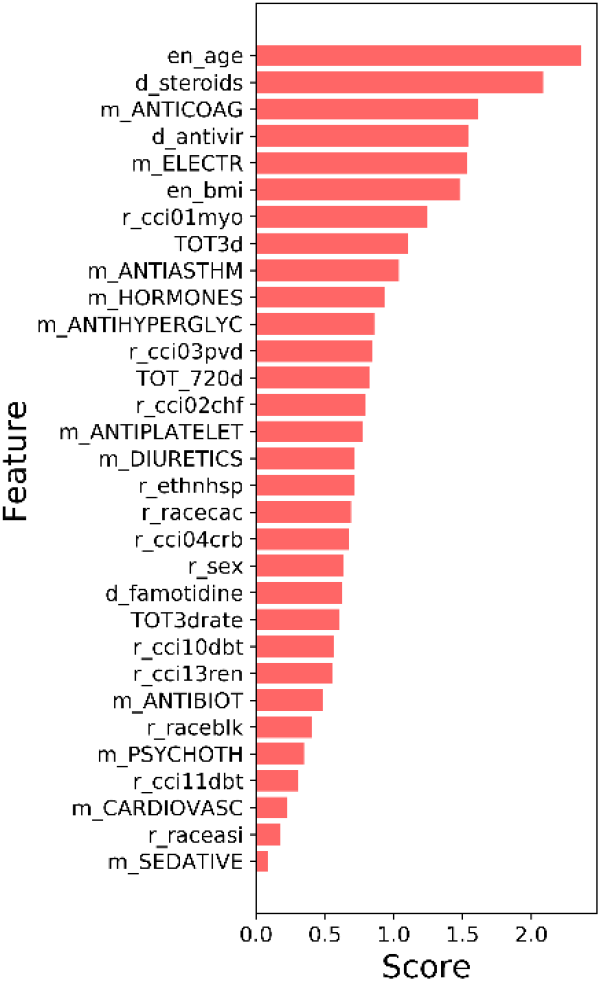
Features scoring for hospitalization stay with DL regression model.

**Figure 5.**
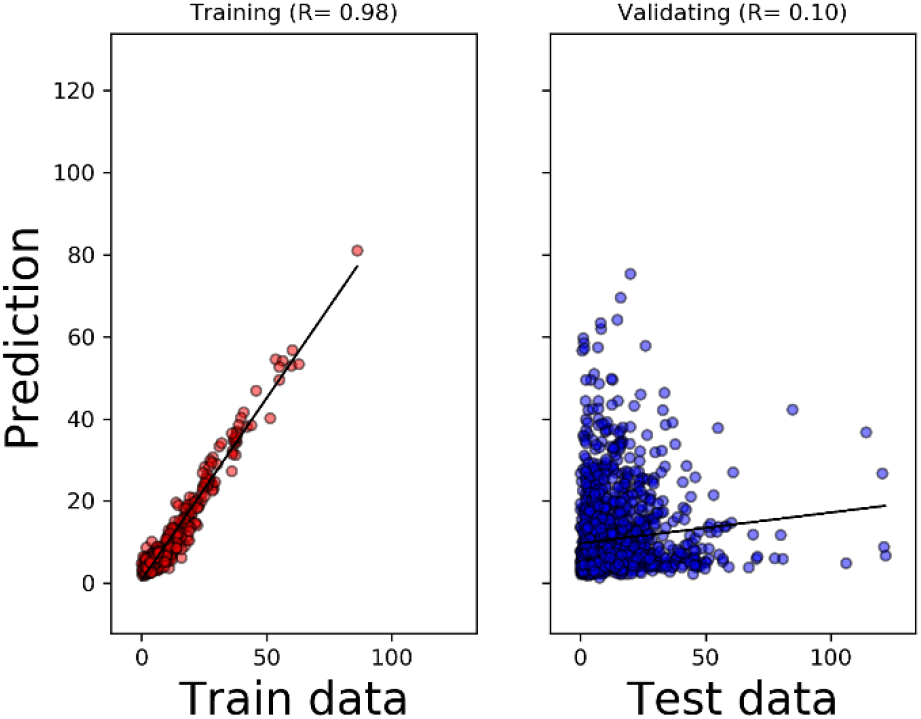
Regression model to predict hospitalization stay indicated further steps needed to optimize the model.

## Data Availability

The raw data sets can be requested by contacting HMH. The data sets generated in this study for the purpose of tables and figures are available upon reasonable request to the authors.

## Acknowledgments

None

## Data availability

The raw data sets can be requested by contacting HMH. The data sets generated in this study for the purpose of tables and figures can be requested directly form the authors.

